# Development and validation of total and regional body composition prediction equations from anthropometry and single frequency segmental bioelectrical impedance with DEXA

**DOI:** 10.1101/2020.12.16.20248330

**Authors:** Richard Powell, Emanuella De Lucia Rolfe, Felix R. Day, John R.B. Perry, Simon J. Griffin, Nita G. Forouhi, Soren Brage, Nicholas J. Wareham, Claudia Langenberg, Ken K. Ong

**Author notes:** **Corresponding author information:** Richard Powell, NIHR Cambridge Biomedical Research Centre – Diet, Anthropometry and Physical Activity Group, MRC Epidemiology Unit, University of Cambridge, Institute of Metabolic Science, Cambridge Biomedical Campus Box 285, Cambridge CB2 0QQ, United Kingdom.

## Abstract

**Aims:** Single-frequency segmental Bioelectrical Impedance Analysis (BIA) is commonly used to estimate body composition. To enhance the value of information derived from BIA, especially for use in large-scale epidemiological studies, we developed and validated equations to predict total and regional (arms, legs, trunk, android, gynoid, visceral) body composition parameters (lean mass and fat mass) from anthropometry and single-frequency (50 kHz) segmental BIA variables, using Dual Energy X-ray Absorptiometry (DEXA) as the criterion method.

**Methods:** The 11,559 adults (age 30 to 65) from the UK population-based Fenland Study with data on DEXA, BIA and anthropometry were randomly assigned to a Derivation sample (4,827 men; 5,732 women) or a Validation sample (500 men; 500 women). Prediction equations based on anthropometry and BIA variables were derived using forward stepwise multiple linear regression in the Fenland Derivation sample. These were validated in the Fenland Validation sample and also in the UK Biobank Imaging Study (2,392 men; 2,606 women) using Pearson correlations and Bland–Altman models.

**Results and Conclusions:** Bland Altman analyses revealed no significant mean bias for any predicted DEXA parameter (all P>0.05) for the fenland population. Bias expressed as % of the mean was between -0.6% and 0.5% for all parameters in both men and women, except for visceral FM and subcutaneous abdominal FM (range -3.6 to 1.1%). However, in UK Biobank most predicted parameters showed significant bias: % mean bias was <2% in both sexes only for total fat mass and total lean mass, and was >10% for leg and visceral fat mass in both sexes. In conclusion, new equations based on anthropometry and BIA variables predicted DEXA parameters with sufficient accuracy to assess relative differences between individuals, and were sufficiently accurate to predict absolute values for total body but not regional fat and lean mass.

## INTRODUCTION

The prevalence of overweight and obesity continues to increase causing a global health problem^1^. Beyond simple anthropometric parameters of obesity (i.e. body mass index and waist circumference), body composition is an important component of health. In particular, the total amount and regional distribution of fat and lean tissues are important risk factors for disease^2-4^. Dual Energy X-ray Absorptiometry (DEXA) and single-frequency segmental Bioelectrical Impedance Analysis (BIA) are methods to estimate total and regional body composition in epidemiological studies^5-6^.

DEXA is a common reference method used to validate other body composition methods^7,8^. DEXA estimates body composition by the attenuation of X-rays at two energies as they pass through body tissues providing estimates of total body and regional body composition for fat mass (FM), lean mass (LM) and bone mass (BM). FM measured by DEXA Lunar Prodigy or iDEXA was shown to correlate highly with the gold standard four component method (4-C) (both: r=0.99) although there was significant mean bias (Lunar Prodigy: -2.16kg; iDEXA: -0.94kg)^9^.

BIA estimates body composition from the impedance of an electrical current, together with other anthropometric data to predict body composition. Single-frequency segmental BIA uses 8 electrodes at 50kHz to provide estimates of total body and regional body composition by incorporating data on other variables, such as height, weight, age, gender and impedance index (height^2^/impedance)^10,11^. Unfortunately BIA manufacturers do not provide the equations used to derive total body and regional body composition estimates^12^. Furthermore, the populations in whom such prediction models are derived and validated should be similar to those in whom they will be applied^8,11^.

There is limited literature on the performance of single-frequency segmental BIA, and most of those studies validated only BIA manufacturer-predicted total body composition values^12,13^. Furthermore, most reported BIA prediction models are generated in small studies, often with specific demographic characteristics^10,11^. Pietrobelli et al^13^ reported high correlation (r=0.89) but statistically significant differences between single-frequency segmental BIA (Tanita BC-418) and DEXA (DPX Lunar) estimates for % total body FM (BIA 27.7±9.2%; DEXA 29.2±10.7%; mean bias 1.5%) and also for regional % fat estimates (arms: r=0.79 to 0.8, bias -2.9% to -3.8%; legs: r=0.8 to 0.85, bias -0.1% to 0.1%, trunk & head: r=0.83, bias 3.7%) (N=40, 50% male, age 28.6±18.3; BMI 24.8±6.1). Similarly, in a Taiwanese population, Chen et al^14^ reported high correlation (r=0.916) between BIA (Tanita BC-418) and DEXA (GE Lunar Prodigy) estimates of total body % FM with significant mean bias (−3.72%) (N=711 58% male, age 34.9±16, BMI 24.4±4.1).

Therefore, we aimed to develop equations for the prediction of total and regional FM and LM in adults from anthropometry and single frequency segmental BIA variables, by comparison against DEXA as the criterion method. We derived these equations in the UK Fenland Study (Derivation Sample) and validated them in independent samples (Fenland Validation Sample & the UK Biobank Imaging study), which had the DEXA and segmental BIA measures.

## METHODS

### The Fenland Study

New prediction models were derived and then validated in separate samples of the Fenland Study (DOI: 10.22025/2017.10.101.00001), a population-based cohort of adults recruited from general practice lists in Cambridgeshire, (Cambridge, Ely, Wisbech, and surrounding villages) in the UK^5^. In total 12,435 individuals (97.6% of European descent) born between 1950-1975 (age 29-65 years at recruitment) attended the baseline clinical examination in 2005-2015. This study was established to investigate the environmental and genetic risk factors for obesity and related co-morbidities. The study was approved by the Cambridge Local Research Ethics Committee (04/Q0108/19), and all participants gave written informed consent.

For the current analysis, we excluded individuals if essential measurements (age, height, weight, BIA and DEXA) were missing (n=622), if tissue was omitted from the DEXA scan (n=184), or if their BIA values were biologically implausible (n=70, >4 SD beyond the mean). The excluded participants were younger and had higher BMI values than the included participants (supplementary table 1). After exclusions, this analysis included 11,559 individuals (5,327 men and 6,232 women). We randomly divided the included participants into a ‘Derivation’ sample (4,827 men; 5,732 women) and a ‘Validation’ sample (500 men; 500 women) see table 1.

**Table 1:** Characteristics of Fenland Derivation and Validation samples and the UK Biobank Imaging Study

| (Mean ± SD) | Derivation Group |  | Validation Group |  | UK BioBank Imaging study |  |
| --- | --- | --- | --- | --- | --- | --- |
|  | Men | Women | Men | Women | Men | Women |
|  | (n=4,827) | (n=5,732) | (n=500) | (n=500) | (n= 2,392) | (n= 2,606) |
| Age (years) | 48.6 ± 7.6 | 48.6 ± 7.4 | 48.5 ± 7.5 | 48.8 ± 7.4 | 62.6* ± 7.5* | 61.2* ± 7.5* |
| Height (cm) | 177.5 ± 6.6 | 164.0 ± 6.3 | 177.2 ± 6.4 | 164.0 ± 6.5 | 176* ± 6.6* | 162.5* ± 6.3* |
| Weight (kg) | 85.6 ± 13.2 | 70.7 ± 13.8 | 85.1 ± 13.4 | 71.0 ± 13.4 | 84.6* ± 14* | 69.7* ± 13.3* |
| BMI (kg/m<sup>2</sup>) | 27.2 ± 3.9 | 26.3 ± 4.9 | 27.1 ± 3.9 | 26.4 ± 4.9 | 27.3* ± 4.0 | 26.4* ± 4.9 |
| Impedance (ohms) | 553.2 ± 57.4 | 672.2 ± 72.0 | 554.3 ± 57.9 | 670.7 ± 72.0 | 545.8* ± 58.6* | 664.4* ± 73.9* |
| Right Leg Impedance (ohms) | 243.4 ± 27.7 | 276.2 ± 33.2 | 244.3 ± 29.5 | 276.0 ± 32.7 | 236.3* ± 30.8* | 273* ± 36.1* |
| Left Leg Impedance (ohms) | 245.2 ± 27.8 | 276.5 ± 33.2 | 246.1 ± 29.2 | 275.8 ± 32.6 | 237.9* ± 30.3* | 273.1* ± 35.8* |
| Right Arm Impedance (ohms) | 284.7 ± 31.3 | 366.7 ± 41.1 | 284.0 ± 30.9 | 366.4 ± 40.7 | 281.5* ± 31.9* | 357.2* ± 40.5* |
| Left Arm Impedance (ohms) | 289.9 ± 32.8 | 374.4 ± 42.5 | 290.2 ± 32.4 | 373.7 ± 42.0 | 286.7* ± 32.1* | 365.8* ± 42.4* |
| Average Waist (cm) | 96.7 ± 11.0 | 85.2 ± 12.1 | 96.4 ± 11.0 | 85.7 ± 12.0 | 93.8* ± 10.1* | 82.2* ± 11.6* |
| Average Hip (cm) | 102.9 ± 6.6 | 103.1 ± 9.8 | 102.7 ± 6.8 | 103.4 ± 9.4 | 101.9* ± 7.4* | 101.3* ± 10* |
| Waist Hip Ratio | 0.9 ± 0.1 | 0.8 ± 0.1 | 0.9 ± 0.1 | 0.8 ± 0.1 | 0.9* ± 0.1* | 0.8* ± 0.1* |
| DEXA Total Bone Mass (g) | 3144.0 ± 382.8 | 2389.0 ± 306.2 | 3130.3 ± 399.2 | 2377.1 ± 313.1 | 3084.7* ± 416.3* | 2230* ± 334.1* |
| DEXA Total Fat Mass (g) | 25209.5 ± 8141.3 | 27131.0 ± 9770.6 | 25011.0 ± 8317.7 | 27344.9 ± 9453.0 | 24988.9 ± 8977.7 | 26819.5 ± 9623.8 |
| DEXA Total Lean Mass (g) | 57192.2 ± 6735.1 | 41310.9 ± 5267.5 | 56983.5 ± 7127.8 | 41343.3 ± 5317.5 | 55583.9* ± 6681.3* | 39731.8* ± 4782.7* |
| DEXA Arms Bone Mass (g) | 448.6 ± 59.9 | 298.0 ± 39.4 | 447.5 ± 61.3 | 295.2 ± 40.4 | 451.4* ± 69.0 | 282.7* ± 53* |
| DEXA Arms Fat Mass (g) | 2357.5 ± 708.0 | 2890.9 ± 928.5 | 2337.5 ± 707.6 | 2923.8 ± 882.3 | 2371.7* ± 798.1 | 2924.8* ± 1049.3 |
| DEXA Arms Lean Mass (g) | 6804.1 ± 1098.8 | 3930.4 ± 701.8 | 6796.3 ± 1134.7 | 3918.8 ± 725.0 | 6781* ± 1054.3 | 4099.3* ± 651.4* |
| DEXA Trunk Bone Mass (g) | 946.6 ± 146.9 | 721.2 ± 120.6 | 940.6 ± 154.7 | 714.9 ± 121.1 | 914* ± 175.5* | 650.4* ± 133.9* |
| DEXA Trunk Fat Mass (g) | 14448.8 ± 5483.9 | 13289.4 ± 5970.9 | 14330.2 ± 5613.9 | 13464.4 ± 5963.4 | 15356.5* ± 6168.4* | 13991.8* ± 6045.8* |
| DEXA Trunk Lean Mass (g) | 26896.3 ± 3198.3 | 20396.8 ± 2523.9 | 26749.0 ± 3359.2 | 20449.5 ± 2588.1 | 26461.7* ± 3110.1* | 19455.4* ± 2274.6* |
| DEXA Legs Bone Mass (g) | 1204.6 ± 165.9 | 853.4 ± 121.4 | 1197.5 ± 169.1 | 853.6 ± 123.4 | 1176.7* ± 168.4* | 804.8* ± 126.8* |
| DEXA Legs Fat Mass (g) | 7453.5 ± 2299.3 | 10154.8 ± 3411.4 | 7397.4 ± 2289.2 | 10161.2 ± 3140.6 | 6298.2* ± 2326.9* | 9091.7* ± 3129.1* |
| DEXA Legs Lean Mass (g) | 20143.5 ± 2761.6 | 14208.6 ± 2241.2 | 20097.0 ± 2913.1 | 14208.1 ± 2187.3 | 18910.3* ± 2773.4* | 13305.9* ± 2062.2* |
| DEXA Android Bone Mass (g) | 53.9 ± 11.1 | 45.9 ± 9.2 | 53.4 ± 11.2 | 45.5 ± 9.1 | 56.2* ± 12.8* | 42.4* ± 9.5* |
| DEXA Android Fat Mass (g) | 2536.5 ± 1151.7 | 2120.8 ± 1163.0 | 2513.7 ± 1160.8 | 2159.2 ± 1171.4 | 2710.3* ± 1240.4* | 2216.4* ± 1157.5* |
| DEXA Android Lean Mass (g) | 4255.6 ± 546.9 | 3139.8 ± 437.9 | 4230.9 ± 557.8 | 3157.4 ± 437.3 | 4099.2* ± 541.4* | 2910.2* ± 415.9* |
| DEXA Gynoid Bone Mass (g) | 321.1 ± 50.1 | 234.5 ± 36.9 | 318.7 ± 52.4 | 234.8 ± 37.9 | 326.1* ± 52.5* | 226.4* ± 37.4* |
| DEXA Gynoid Fat Mass (g) | 3710.7 ± 1188.2 | 4957.5 ± 1601.0 | 3687.2 ± 1225.9 | 4950.3 ± 1506.3 | 3596.3* ± 1317.7* | 4683.7* ± 1551.2* |
| DEXA Gynoid Lean Mass (g) | 9130.6 ± 1185.7 | 6542.1 ± 890.5 | 9078.9 ± 1218.8 | 6538.8 ± 912.2 | 8630.2* ± 1174.6* | 6125.6* ± 800.5* |
| DEXA VAT (g) | 1348.1 ± 816.4 | 598.4 ± 534.8 | 1349.3 ± 817.9 | 638.1 ± 578.8 | 1695.2* ± 945.2* | 777.6* ± 583.3* |
| DEXA SCAT (g) | 1189.8 ± 536.2 | 1523.1 ± 729.7 | 1167.1 ± 548.4 | 1521.1 ± 689.5 | 993.3* ± 457.2* | 1425.7* ± 656.1* |
| DEXA Body Fat (%) | 28.9 ± 5.9 | 37.3 ± 6.9 | 28.8 ± 6.0 | 37.6 ± 6.8 | 30.3* ± 6.5* | 39.2* ± 7.3* |

\*p<0.05 from Students t-test compared to the Fenland Derivation Sample

### UK Biobank Imaging Study

The prediction equations were further validated in the UK Biobank Imaging Study. UK Biobank is a population-based cohort study of ∼500,000 individuals aged between 40-69 years at baseline, who were recruited in 2006-2010 from several centres across the United Kingdom. Adults were re-invited to attend the UK Biobank Imaging Study in 2014, including 5,112 participants with whole body iDEXA scans. The sample included here comprised 2,392 men and 2,606 women with biologically plausible measurements of anthropometry, DEXA, and segmental BIA (Table 1).

UK Biobank received approval from the National Information Governance Board for Health and Social Care and the National Health Service North West Centre for Research Ethics Committee (Ref: 11/NW/0382)^15^.

### Anthropometry

In both studies, anthropometry and body composition measurements were performed by trained research staff, following standard protocols^16-18^. Body weight and height were measured with the participants barefoot and in light indoor clothes. BMI was calculated as weight (kg) divided by height squared (m^2^).

In the Fenland study^16^, waist circumference was measured at the midpoint between the lower costal margins and the level of the anterior superior iliac crests, and hip circumference was measured at the widest level of the greater trochanters. Waist and hip circumferences were measured using a non-stretchable fibre-glass tape. In UK Biobank, waist circumference was measured at the level of the smallest part of the trunk or the umbilicus, and hip circumference was measured at the level of the widest part of the hips. Waist and hip circumferences were measured using a SECA 200 tape measure^14^.

### Segmental BIA

In both studies, BIA was performed using the Tanita BC-418MA Segmental Body Composition Analyser (Tanita Corporation) adhering to the manufacturer’s guidelines. This device is a single-frequency (50kHz) BIA monitor that uses eight polar electrodes and a single-point load cell weighing system in the scale platform. It provides separate body mass and impedance readings for different body segments, such as right arm, left arm, right leg, and left leg. For the current analysis, we summed left and right limb values and calculated impedance indices (height^2^ in m^2^ / impedance in ohms) for the whole body, arms and legs.

### DEXA

In the Fenland study, most whole body DEXA scans were performed using Lunar Prodigy Advanced (GE Healthcare), and a minority (2.2%) used iDEXA (GE Healthcare). In the UK Biobank Imaging Study, all whole body DEXA scans were performed using iDEXA. Participants were scanned by trained operators using standard imaging and positioning protocols. Before scanning, DEXA systems were calibrated according to the manufacturer’s guidelines using a spine phantom made of calcium hydroxyapatite, embedded in a lucite block (GE-Lunar, Madison, WI)^16,18^.

In both studies, and for data from both DEXA scanners in the Fenland study, enCORE software version 14-16 (GE Healthcare) was used under the enhanced analysis protocol to acquire total and regional FM and LM. The enCORE software automatically demarcated the boundaries of body regions which were checked and adjusted where needed by trained operators^15^.

### Statistical analyses

Statistical analyses were performed using STATA version 16.1 (StataCorp, College Station, TX). Descriptive data are reported as means ± SD, P value <0.05 was considered statistically significant.

The performance of these different models was compared by calculating the explained variance in each outcome parameter (model Pearson correlation coefficients) and root mean square deviation (RMSD) values, which represent the average deviance between the observed (measured) and predicted values. Derivation of prediction models: in the Fenland Derivation sample, sex-stratified forward stepwise (p<0.05) regression models were performed separately for the following dependent variables: total body and regional FM and LM DEXA parameters. We note that the chosen DEXA parameters are based on a 3-compartment model, in which FM and LM are distinct from bone mass. Collinearity between co-variates was indicated by model mean variance inflation factor >10; by this criterion, waist-hip ratio, total body impedance index, and individual segmental impedance values were removed. For each outcome, four different models were compared:

> Basic model: (age, height and weight)
>
> Model A: (age, height, weight, waist and hip circumferences)
>
> Model B: (age, height, weight, total body impedance, arm and leg impedance indices)
>
> Model C: (age, height, weight, waist and hip circumferences, total body impedance, arm and leg impedance indices)

The performance of these different models was compared by calculating the explained variance in each outcome parameter (model r-square values) and root mean square deviation (RMSD) values, which represent the average deviance between the observed and regression model predicted values.

### Model validation

In the Fenland Validation sample and the UK Biobank Imaging Study, total body and regional FM and LM values were predicted in each individual using the final model equations. The level of agreement to the observed measure was assessed using: Pearson’s correlations. With Student’s t-test and linear regression analysis used to identify any significant differences in the point estimate. Bland-Altman plots were also used to assess the agreement, and the root mean square error (RMSE) was calculated. Bias was calculated as the difference between equation predicted and DEXA measured values, and the limits of agreement as the 95% confidence range (mean bias +1.96 standard deviations).

## RESULTS

The characteristics of the Fenland Derivation, Validation samples and the UK Biobank Imaging Study are summarised in Table 2. There was no significant difference between the Fenland Derivation and Validation samples in any parameter. Compared to the Fenland Derivation sample, individuals in the UK Biobank Imaging Study had similar BMI values but were older, shorter, lighter, and had lower BIA and DEXA parameters.

**Table 2:**

|  |  |
| --- | --- |
| *Total Fat Mass (in grams) in men =* | -15486 + (48.98*Age in years) - (184.9*Height in cm) +<br>(590*Weight in kg) + (153.1*Waist in cm) + (71.28*Hip in<br>cm) + (21.7*Total Impedance in ohm) - (467222*Legs<br>impedance index) - (1881000*Arms impedance index) |
| Arms impedance index ($\text{height}^2$ in $\text{m}^2$ / left and right arm impedance in ohm) | |
| Legs impedance index ($\text{height}^2$ in $\text{m}^2$ / left and right leg impedance in ohm) | |

### Derivation of prediction models

Comparing the results of multiple linear regression models, the most comprehensive model, i.e. Model C (including age, height, weight, waist and hip circumferences, total body impedance, arm and leg impedance indices), consistently produced higher r-square values and lower RMSD values, compared to the other models (Supplementary Table 2). Thus this was the basis of the final prediction models. In these the weights were built based on the beta coefficients from Model C, as were the constant values, shown in Supplementary Table 3. For example:

### Validation of prediction models in the Fenland Study

In the Fenland Validation sample, DEXA total body and regional FM and LM parameters were predicted from anthropometry and BIA variables using the equations derived above from Model C (Supplementary Table 2). Correlation coefficients between predicted and measured DEXA parameters were strong (R^2^ >0.8) for all FM and LM variables; the minimum was for subcutaneous abdominal FM in men. (Table 3). Bland Altman analyses revealed no significant mean bias for any predicted DEXA parameter (all P>0.05). Bias expressed as % of the mean was between -0.6% and 0.5% for all parameters in both men and women, except for visceral FM and subcutaneous abdominal FM (range -3.6 to 1.1%).

**Table 3:** Agreement between predicted and measured DEXA parameters (all in grams) in the Fenland Validation sample. Pearson’s correlation coefficients, and mean bias are shown.

|  | Men |  |  |  |  |
| --- | --- | --- | --- | --- | --- |
| | r | Mean Bias ± 95% range | Bias % mean | RMSE % mean | P* |
| DEXA Total Fat Mass (g) | 0.96 | 20.2 ± 4675.0 | 0.1 | 9.5 | 0.85 |
| DEXA Total Lean Mass (g) | 0.94 | 3.7 ± 4676.0 | 0.0 | 4.2 | 0.97 |
| DEXA Arms Fat Mass (g) | 0.88 | -4.4 ± 673.9 | -0.2 | 14.6 | 0.78 |
| DEXA Arms Lean Mass (g) | 0.89 | 10.2 ± 1055.3 | 0.2 | 7.9 | 0.67 |
| DEXA Trunk Fat Mass (g) | 0.96 | 21.8 ± 3337.2 | 0.2 | 11.8 | 0.78 |
| DEXA Trunk Lean Mass (g) | 0.88 | -48.8 ± 2957.5 | -0.2 | 5.6 | 0.47 |
| DEXA Legs Fat Mass (g) | 0.87 | 4.2 ± 2157.5 | 0.1 | 14.8 | 0.93 |
| DEXA Legs Lean Mass (g) | 0.91 | 42.1 ± 2293.7 | 0.2 | 5.8 | 0.42 |
| DEXA Android Fat Mass (g) | 0.95 | 4.8 ± 720.1 | 0.2 | 14.5 | 0.77 |
| DEXA Android Lean Mass (g) | 0.85 | -7.5 ± 575.6 | -0.2 | 6.9 | 0.57 |
| DEXA Gynoid Fat Mass (g) | 0.91 | 6.1 ± 967.6 | 0.2 | 13.3 | 0.78 |
| DEXA Gynoid Lean Mass (g) | 0.89 | -20.0 ± 1048.0 | -0.2 | 5.9 | 0.40 |
| DEXA VAT (g) | 0.87 | 23.7 ± 786.0 | 1.8 | 29.7 | 0.19 |
| DEXA SCAT (g) | 0.80 | -12.1 ± 672.4 | -1.0 | 28.8 | 0.43 |

|  | Women |  |  |  |  |
| --- | --- | --- | --- | --- | --- |
| | r | Mean Bias ± 95% range | Bias % mean | RMSE % mean | P* |
| DEXA Total Fat Mass (g) | 0.98 | 42.4 ± 3554.9 | 0.2 | 6.7 | 0.60 |
| DEXA Total Lean Mass (g) | 0.94 | -18.6 ± 3531.6 | 0.0 | 4.4 | 0.82 |
| DEXA Arms Fat Mass (g) | 0.91 | 17.9 ± 721.4 | 0.6 | 12.7 | 0.28 |
| DEXA Arms Lean Mass (g) | 0.87 | -20.4 ± 698.9 | -0.5 | 9.1 | 0.20 |
| DEXA Trunk Fat Mass (g) | 0.97 | 20.2 ± 2697.4 | 0.2 | 10.4 | 0.74 |
| DEXA Trunk Lean Mass (g) | 0.87 | 26.4 ± 2479.9 | 0.1 | 6.2 | 0.64 |
| DEXA Legs Fat Mass (g) | 0.92 | 6.1 ± 2476.0 | 0.1 | 12.4 | 0.91 |
| DEXA Legs Lean Mass (g) | 0.94 | -16.2 ± 1517.0 | -0.1 | 5.4 | 0.64 |
| DEXA Android Fat Mass (g) | 0.96 | 5.8 ± 634.3 | 0.3 | 15.3 | 0.69 |
| DEXA Android Lean Mass (g) | 0.84 | 11.3 ± 466.1 | 0.4 | 7.6 | 0.29 |
| DEXA Gynoid Fat Mass (g) | 0.95 | -15.7 ± 954.1 | -0.3 | 9.8 | 0.47 |
| DEXA Gynoid Lean Mass (g) | 0.90 | -6.2 ± 768.0 | -0.1 | 6.0 | 0.72 |
| DEXA VAT (g) | 0.89 | 21.3 ± 527.1 | 3.6 | 44.9 | 0.08 |
| DEXA SCAT (g) | 0.92 | -17.3 ± 540.1 | -1.1 | 18.1 | 0.16 |

r, Pearson's correlation coefficients (all correlations were $P < 0.0001$ )
\* t-test of the difference between the point estimate of the mean difference and zero
Bias = predicted value - reference value

**Table 4:** Agreement between predicted and measured DEXA parameters (all in grams) in the UK Biobank Imaging Study. Pearson’s correlation coefficients, and mean bias are shown.

|  | Men |  |  |  |  |
| --- | --- | --- | --- | --- | --- |
| | r | Mean Bias ± 95% range | Bias % mean | RMSE % mean | P* |
| DEXA Total Fat Mass (g) | 0.97 | 59.9 ± 4839.1 | 0.2 | 9.8 | 0.24 |
| DEXA Total Lean Mass (g) | 0.93 | -894.8 ± 4965.6 | -1.6 | 4.4 | 0.00 |
| DEXA Arms Fat Mass (g) | 0.89 | -31.9 ± 722.2 | -1.4 | 15.6 | 0.00 |
| DEXA Arms Lean Mass (g) | 0.88 | 204.1 ± 1042.0 | 3.0 | 7.8 | 0.00 |
| DEXA Trunk Fat Mass (g) | 0.96 | 1024.6 ± 3848.1 | 7.1 | 13.6 | 0.00 |
| DEXA Trunk Lean Mass (g) | 0.84 | -617.4 ± 3373.0 | -2.3 | 6.4 | 0.00 |
| DEXA Legs Fat Mass (g) | 0.88 | -942.3 ± 2211.2 | -12.6 | 15.1 | 0.00 |
| DEXA Legs Lean Mass (g) | 0.92 | -603.9 ± 2125.8 | -3.0 | 5.4 | 0.00 |
| DEXA Android Fat Mass (g) | 0.94 | 184.4 ± 835.9 | 7.3 | 16.8 | 0.00 |
| DEXA Android Lean Mass (g) | 0.82 | -196.3 ± 617.6 | -4.6 | 7.4 | 0.00 |
| DEXA Gynoid Fat Mass (g) | 0.92 | 3.1 ± 999.3 | 0.1 | 13.7 | 0.77 |
| DEXA Gynoid Lean Mass (g) | 0.89 | -333.6 ± 1077.8 | -3.7 | 6.0 | 0.00 |
| DEXA VAT (g) | 0.86 | 223.5 ± 1014.5 | 16.6 | 38.4 | 0.00 |
| DEXA SCAT (g) | 0.72 | -38.9 ± 647.1 | -3.3 | 27.8 | 0.00 |

|  | Women |  |  |  |  |
| --- | --- | --- | --- | --- | --- |
| | r | Mean Bias ± 95% range | Bias % mean | RMSE % mean | P* |
| DEXA Total Fat Mass (g) | 0.98 | -371.2 ± 3389.5 | -1.4 | 6.4 | 0.00 |
| DEXA Total Lean Mass (g) | 0.93 | -492.7 ± 3528.1 | -1.2 | 4.4 | 0.00 |
| DEXA Arms Fat Mass (g) | 0.89 | -75.8 ± 956.7 | -2.6 | 16.9 | 0.00 |
| DEXA Arms Lean Mass (g) | 0.85 | 242.8 ± 673.5 | 6.2 | 8.7 | 0.00 |
| DEXA Trunk Fat Mass (g) | 0.96 | 662.3 ± 3275.3 | 5.0 | 12.6 | 0.00 |
| DEXA Trunk Lean Mass (g) | 0.83 | -618.0 ± 2526.2 | -3.0 | 6.3 | 0.00 |
| DEXA Legs Fat Mass (g) | 0.91 | -990.8 ± 2574.3 | -9.8 | 12.9 | 0.00 |
| DEXA Legs Lean Mass (g) | 0.93 | -236.0 ± 1531.2 | -1.7 | 5.5 | 0.00 |
| DEXA Android Fat Mass (g) | 0.95 | 118.3 ± 700.4 | 5.6 | 16.8 | 0.00 |
| DEXA Android Lean Mass (g) | 0.83 | -169.4 ± 460.3 | -5.4 | 7.5 | 0.00 |
| DEXA Gynoid Fat Mass (g) | 0.95 | -203.0 ± 992.4 | -4.1 | 10.2 | 0.00 |
| DEXA Gynoid Lean Mass (g) | 0.90 | -195.8 ± 706.5 | -3.0 | 5.5 | 0.00 |
| DEXA VAT (g) | 0.86 | 95.1 ± 606.8 | 15.9 | 51.7 | 0.00 |
| DEXA SCAT (g) | 0.91 | 18.0 ± 535.7 | 1.2 | 17.9 | 0.00 |

r, Pearson's correlation coefficients (all correlations were $P < 0.0001$ )
\* t-test of the difference between the point estimate of the mean difference and zero
Bias = predicted value - reference value

### Validation of prediction models in the UK Biobank Imaging Study

In the UK Biobank Imaging Study, DEXA total body and regional FM and LM parameters were predicted from anthropometry and BIA variables using the equations derived above from Model C (Supplementary Table 2). Correlation coefficients between predicted and measured DEXA parameters were strong for all FM and LM variables; again, the minimum was for subcutaneous abdominal FM in men (r=0.72).

However, Bland Altman analyses revealed significant (P<0.05) mean bias for all predicted DEXA parameters with the exception of total FM and gynoid FM in men. Bias expressed as % of the mean for each parameter was between -0.2% and 1.6% for total FM and total LM in both sexes. For most other parameters, % mean bias was between -7.5% and 5.5% and was >9.5% for leg and visceral FM in both sexes.

## DISCUSSION

We developed and validated equations to predict total and regional DEXA FM and LM parameters from anthropometry and BIA values, separately in men and women. In both independent study samples, we found that all predicted parameters showed high correlations with DEXA measured parameters. All parameters showed Pearson correlation coefficients >0.7, and the large majority showed correlations of >0.85. This provides sufficient accuracy to be used in future studies that aim to analyse relative differences between individuals, for example to identify determinants of the continuous distribution in body composition parameters. Our predictions of % total body fat (Fenland Validation: r=0.92, mean bias 0.03%; UK Biobank: r=0.94, 0.05%) are better than those reported using manufacturer-predicted BIA values using the Tanita BC-418 (r=0.89, 1.5%)^12^.

In the Fenland Validation sample, who were drawn from the same population and were similar in all characteristics to the Fenland Derivation sample, the predicted parameters showed modest and non-significant mean bias, mostly between -1 and 1%. However in the UK Biobank sample, our predicted values showed significant bias with the exception of total body fat mass and gynoid fat mass in men. These difference may relate at least in part to the differences in DEXA machines used between the 2 studies; it was previously reported that the Lunar Prodigy overestimates FM (mean bias 1.18kg) and underestimates LM (mean bias -1.29kg) compared to the iDEXA^5^. Furthermore, UK Biobank participants were on average 12 to 14 years older and slightly shorter and lighter than those in the Fenland Validation sample. Therefore, caution is needed if using these equations to predict absolute values for regional body composition, for example to identify determinants of categories of body composition above or below a specific absolute value.

Strengths of our study include the large sizes of both the Fenland and UK Biobank studies, the use of the same BIA models and the same DEXA analytical software, and robust validation in two separate independent samples. We acknowledge that our study samples were predominantly of white Caucasian origin. Future studies should assess the validity of these equations in populations from other ethnic groups. While DEXA is widely accepted as a criterion method for most total and regional FM and LM quantities, neither DEXA nor BIA are designed to distinguish between superficial and deeper tissues, i.e. between subcutaneous abdominal and visceral FM. Indeed, the weakest agreements were seen for visceral FM and we note that other equations have been derived for this parameter in UK Biobank Imaging study using Tanita BC-418 and iDEXA (n=4,198 r=0.87; 95% CI of 740-780g with a bias between -0.4 and 0.54)^19^. We also acknowledge that DEXA estimates LM distinct from bone mass. By contrast, BIA assumes a 2-compartment model and typically estimates fat-free mass (which includes LM and bone mass). We therefore did not aim to use BIA data to estimate DEXA bone mass parameters.

In summary, BIA is a simple method to assess body composition which is used widely, particularly in very large-scale studies. These new equations enhance the value of information derived from single frequency segmental BIA.

## Data Availability

The processes that relate to access to the Fenland study data are detailed at https://epi-meta.mrc-epid.cam.ac.uk/. All the data used in this study form the UK Biobank resource is available to bona fide researchers for health-related research in the public interest. Researchers who wish to access the data should contact UK Biobank for further details about access to this resource. Further details are available at https://www.ukbiobank.ac.uk/enable-your-research/register.

## Acknowledgments

This research has been conducted using data from the Fenland Study and the UK Biobank Resource (project ID 44448). We are grateful to all study participants and the research teams responsible for data collection, and to the following funding bodies:

The Fenland Study was funded by the Medical Research Council and the Wellcome Trust.

The Biobank Study was funded by the UK Medical Research Council, Wellcome Trust, Department of Health, British Heart Foundation, Diabetes UK, Northwest Regional Development Agency, Scottish Government, and Welsh Assembly Government.

We also acknowledge support from the Medical Research Council (Unit programmes MC_UU_12015/1, MC_UU_12015/2, MC_UU_12015/3, MC_UU_12015/4 and MC_UU_12015/5).

We are grateful to funding to RP and EDLR, who are supported by the NIHR Biomedical Research Centre Cambridge [IS-BRC-1215-20014]. The NIHR Cambridge Biomedical Research Centre (BRC) is a partnership between Cambridge University Hospitals NHS Foundation Trust and the University of Cambridge, funded by the National Institute for Health Research (NIHR). The views expressed are those of the authors and not necessarily those of the NHS, the NIHR or the Department of Health and Social Care.

