## Supplementary tables1-3 for "Development and validation of total and regional body composition prediction equations from anthropometry and single frequency segmental bioelectrical impedance with DEXA"

**Supplementary Table 1**: Characteristics of excluded participants from the Fenland study

| (Mean ± SD) | Men (n=416) | Women (n=458) | Total (n=876) |
|---|---|---|---|
| Age (years) | 47.73 ± 7.79 | 48.17 ± 7.77 | 47.97 ± 7.78 |
| Height (cm) | 179.15 ± 8.06 | 163.56 ± 7.03 | 170.95 ± 10.84 |
| Weight (kg) | 95.46 ± 22.00 | 79.25 ± 24.03 | 86.93 ± 24.46 |
| BMI (kg/m$^2$) | 29.66 ± 6.13 | 29.62 ± 8.68 | 29.64 ± 7.58 |
| Impedance (ohms) | 534.21 ± 73.12 | 656.46 ± 100.62 | 598.16 ± 107.56 |
| DEXA PBF (%) | 31.20 ± 6.29 | 42.98 ± 9.30 | 37.43 ± 9.94 |

**Supplementary Table 2**: Comparison of the overall performance of the different regression models for DEXA parameters, in the Fenland Derivation sample.

| | Basic Model | | | | Model A | | | | Model B | | | | Model C | | | |
|---|---|---|---|---|---|---|---|---|---|---|---|---|---|---|---|---|
| | Men | | Women | | Men | | Women | | Men | | Women | | Men | | Women | |
| | r | % RMSE | r | % RMSE | r | % RMSE | r | % RMSE | r | % RMSE | r | % RMSE | r | % RMSE | r | % RMSE |
| Total Fat Mass (g) | 0.92 | 13.0 | 0.97 | 9.1 | 0.93 | 11.5 | 0.97 | 8.6 | 0.96 | 9.4 | 0.98 | 6.5 | 0.96 | 9.0 | 0.98 | 9.0 |
| Total Lean Mass (g) | 0.88 | 5.7 | 0.88 | 6.0 | 0.90 | 5.0 | 0.90 | 5.6 | 0.94 | 4.1 | 0.94 | 4.2 | 0.94 | 4.0 | 0.94 | 4.0 |
| Arms Fat Mass (g) | 0.83 | 16.9 | 0.90 | 14.2 | 0.84 | 16.3 | 0.90 | 14.1 | 0.88 | 14.5 | 0.91 | 13.0 | 0.88 | 14.5 | 0.92 | 14.5 |
| Arms Lean Mass (g) | 0.75 | 10.7 | 0.78 | 11.2 | 0.79 | 9.9 | 0.81 | 10.6 | 0.89 | 7.4 | 0.88 | 8.5 | 0.89 | 7.2 | 0.88 | 7.2 |
| Trunk Fat Mass (g) | 0.91 | 15.7 | 0.94 | 15.0 | 0.94 | 13.3 | 0.96 | 12.8 | 0.95 | 12.2 | 0.96 | 12.7 | 0.96 | 11.1 | 0.97 | 11.1 |
| Trunk Lean Mass (g) | 0.82 | 6.8 | 0.81 | 7.2 | 0.85 | 6.3 | 0.83 | 7.0 | 0.87 | 5.8 | 0.87 | 6.1 | 0.88 | 5.6 | 0.87 | 5.6 |
| Legs Fat Mass (g) | 0.80 | 18.3 | 0.88 | 16.2 | 0.83 | 17.0 | 0.91 | 13.6 | 0.85 | 16.0 | 0.90 | 15.0 | 0.87 | 15.1 | 0.93 | 15.1 |
| Legs Lean Mass (g) | 0.85 | 7.3 | 0.88 | 7.5 | 0.87 | 6.7 | 0.89 | 7.1 | 0.91 | 5.7 | 0.93 | 5.7 | 0.91 | 5.6 | 0.93 | 5.6 |
| Android Fat Mass (g) | 0.90 | 20.1 | 0.92 | 20.9 | 0.93 | 16.7 | 0.95 | 17.7 | 0.93 | 16.4 | 0.94 | 18.5 | 0.95 | 14.6 | 0.96 | 14.6 |
| Android Lean Mass (g) | 0.80 | 7.6 | 0.80 | 8.4 | 0.81 | 7.5 | 0.80 | 8.3 | 0.84 | 6.9 | 0.84 | 7.5 | 0.84 | 6.9 | 0.84 | 6.9 |
| Gynoid Fat Mass (g) | 0.85 | 16.8 | 0.90 | 13.8 | 0.88 | 15.1 | 0.94 | 10.8 | 0.89 | 14.3 | 0.92 | 12.6 | 0.91 | 13.2 | 0.95 | 13.2 |
| Gynoid Lean Mass (g) | 0.83 | 7.3 | 0.84 | 7.3 | 0.86 | 6.6 | 0.86 | 6.9 | 0.88 | 6.3 | 0.90 | 6.0 | 0.89 | 5.9 | 0.90 | 5.9 |
| VAT (g) | 0.81 | 35.4 | 0.81 | 52.7 | 0.86 | 31.2 | 0.86 | 45.5 | 0.84 | 33.2 | 0.82 | 51.2 | 0.87 | 30.3 | 0.87 | 30.3 |
| SCAT (g) | 0.75 | 29.8 | 0.90 | 20.7 | 0.77 | 28.8 | 0.91 | 19.4 | 0.79 | 27.4 | 0.92 | 18.7 | 0.80 | 27.2 | 0.93 | 27.2 |

Basic model (age, height and weight)
Model A (age, height, weight, waist and hip)
Model B (age, height, weight, impedance, arm impedance index and leg impedance index)
Model C (age, height, weight, waist, hip, impedance, arm impedance index and leg impedance index)
arm impedance index (height m$^2$ / left and right arm impedance)
legs impedance index (height m$^2$ / left and right leg impedance)

**Supplementary Table 3**: Results of regression models for DEXA total and regional body composition parameters (in grams) in men and women in the Fenland Derivation sample. Values are Beta coefficients, unless stated otherwise.

| | Men: Basic Model | | | | | | | | | Women: Basic Model | | | | | | | | |
|---|---|---|---|---|---|---|---|---|---|---|---|---|---|---|---|---|---|---|
| | Age | height | weight | waist | hip | Impedance | Ht2ImLegs | Ht2ImArms | Constant | Age | height | weight | waist | hip | impedance | Ht2ImLegs | Ht2ImArms | Constant |
| Total Fat Mass (g) | 50.5 | -295.8 | 605.2 | | | | | | 23,479 | 78.1 | -292.2 | 708.9 | | | | | | 21106 |
| Total Lean Mass (g) | -57.8 | 268.0 | 374.9 | | | | | | -19,641 | -75.6 | 274.9 | 277.0 | | | | | | -19686 |
| Arms Fat Mass (g) | 5.9 | -26.4 | 47.6 | | | | | | 2,687 | 12.7 | -28.4 | 62.1 | | | | | | 2534 |
| Arms Lean Mass (g) | -14.7 | 17.0 | 57.9 | | | | | | -441 | -7.6 | 16.0 | 36.9 | | | | | | -934.9 |
| Trunk Fat Mass (g) | 57.8 | -238.8 | 406.4 | | | | | | 19,252 | 74.8 | -225.7 | 421.3 | | | | | | 16863 |
| Trunk Lean Mass (g) | 9.1 | 125.8 | 165.5 | | | | | | -10,042 | -19.0 | 138.2 | 117.7 | | | | | | -9670 |
| Legs Fat Mass (g) | -13.0 | -30.3 | 145.8 | | | | | | 997.1 | -9.3 | -38.8 | 221.6 | | | | | | 1294 |
| Legs Lean Mass (g) | -49.8 | 117.2 | 143.1 | | | | | | -10,478 | -46.6 | 110.4 | 118.0 | | | | | | -9968 |
| Android Fat Mass (g) | 15.2 | -53.1 | 83.9 | | | | | | 4,048 | 14.4 | -44.7 | 80.5 | | | | | | 3053 |
| Android Lean Mass (g) | 5.4 | 19.7 | 28.1 | | | | | | -1,906 | -1.7 | 23.4 | 20.1 | | | | | | -2028 |
| Gynoid Fat Mass (g) | -6.5 | -31.2 | 81.7 | | | | | | 2,570 | | -27.1 | 108.0 | | | | | | 1764 |
| Gynoid Lean Mass (g) | -18.1 | 53.7 | 58.3 | | | | | | -4,511 | -16.7 | 50.6 | 42.3 | | | | | | -3942 |
| VAT (g) | 23.0 | -37.3 | 52.0 | | | | | | 2,409 | 13.7 | -22.3 | 31.4 | | | | | | 1369 |
| SCAT (g) | -7.8 | -16.1 | 32.7 | | | | | | 1,628 | | -22.8 | 49.8 | | | | | | 1738 |

| | Men: Model A | | | | | | | | | Women: Model A | | | | | | | | |
|---|---|---|---|---|---|---|---|---|---|---|---|---|---|---|---|---|---|---|
| | Age | height | weight | waist | hip | Impedance | Ht2ImLegs | Ht2ImArms | Constant | Age | height | weight | waist | hip | impedance | Ht2ImLegs | Ht2ImArms | Constant |
| Total Fat Mass (g) | | -130.0 | 268.1 | 322.6 | 159.4 | | | | -22262.0 | 62.8 | -210.0 | 483.1 | 108.6 | 197.2 | | | | -5241 |
| Total Lean Mass (g) | -17.6 | 104.0 | 705.8 | -315.8 | -157.0 | | | | 25894.0 | -60.2 | 194.2 | 496.8 | -109.5 | -187.4 | | | | 5910 |
| Arms Fat Mass (g) | 3.3 | -15.7 | 25.5 | 20.7 | 11.2 | | | | -361.7 | 11.2 | -23.8 | 54.2 | 12.25 | -3.436 | | | | 1731 |
| Arms Lean Mass (g) | -10.6 | -10.5 | 125.1 | -46.7 | -62.2 | | | | 9393.0 | -6.6 | 6.3 | 68.8 | -5.808 | -39.4 | | | | 2914 |
| Trunk Fat Mass (g) | 17.6 | -111.7 | 189.6 | 265.9 | | | | | -8530.0 | 48.7 | -133.4 | 232.6 | 210.9 | 19.42 | | | | -3617 |
| Trunk Lean Mass (g) | 26.4 | 50.6 | 322.0 | -142.1 | -86.9 | | | | 11761.0 | -12.6 | 102.8 | 217.3 | -44.38 | -91.37 | | | | 1987 |
| Legs Fat Mass (g) | -9.8 | | 42.3 | 32.1 | 164.7 | | | | -15734.0 | | -53.2 | 190.0 | -114 | 184.2 | | | | -3863 |
| Legs Lean Mass (g) | -31.5 | 59.4 | 241.9 | -121.1 | | | | | 2166.0 | -39.2 | 77.9 | 197.7 | -55.65 | -48.74 | | | | -884.4 |
| Android Fat Mass (g) | 5.4 | -23.8 | 36.1 | 62.7 | -7.0 | | | | -1941.0 | 8.5 | -24.6 | 40.8 | 47.75 | | | | | -1211 |
| Android Lean Mass (g) | 7.1 | 12.3 | 43.6 | -14.0 | -8.9 | | | | 259.4 | -1.3 | 20.0 | 30.5 | -2.657 | -11.89 | | | | -786.8 |
| Gynoid Fat Mass (g) | -5.5 | -14.2 | 25.2 | 20.7 | 84.8 | | | | -6385.0 | 2.7 | -23.3 | 66.7 | -36.54 | 104.4 | | | | -3730 |
| Gynoid Lean Mass (g) | -7.2 | 25.2 | 98.8 | -64.3 | 25.5 | | | | 149.1 | -12.9 | 36.9 | 71.4 | -30.3 | -5.666 | | | | -768.9 |
| VAT (g) | 14.5 | -16.8 | 26.0 | 47.8 | -27.7 | | | | -369.2 | 10.4 | -13.1 | 18.5 | 28.94 | -16.34 | | | | 160.9 |
| SCAT (g) | -9.3 | -7.0 | 11.1 | 15.8 | 18.6 | | | | -1516.0 | -1.9 | -11.3 | 21.2 | 18.61 | 19.04 | | | | -1578 |

| | Men: Model B | | | | | | | | | Women: Model B | | | | | | | | |
|---|---|---|---|---|---|---|---|---|---|---|---|---|---|---|---|---|---|---|
| | Age | height | weight | waist | hip | Impedance | Ht2ImLegs | Ht2ImArms | Constant | Age | height | weight | waist | hip | impedance | Ht2ImLegs | Ht2ImArms | Constant |
| Total Fat Mass (g) | 71.85 | -224.5 | 771.8 | | | 19.61 | -2409000 | -828,983 | 3,485 | 76.9 | -236.6 | 825.5 | | | 14.72 | -1867000 | -960472 | 5509 |
| Total Lean Mass (g) | -79.31 | 204.1 | 208.3 | | | -20.84 | 2264000 | 846,222 | 431 | -74.3 | 231.8 | 161.5 | | | -16.13 | 1604000 | 928206 | -4150 |
| Arms Fat Mass (g) | 7.889 | -23.08 | 62.65 | | | 2.284 | -168536 | -72,640 | 856.1 | 12.5 | -26.9 | 73.4 | | | 1.909 | | -153495 | 983.1 |
| Arms Lean Mass (g) | -17.35 | | 28.61 | | | -2.614 | 1081000 | -320,194 | 2,739 | -7.8 | | 21.5 | | | -1.011 | 797217 | -112363 | 1089 |
| Trunk Fat Mass (g) | 72.08 | -190.4 | 512.8 | | | 12.12 | -1227000 | -814,959 | 6,308 | 73.5 | -201.9 | 494.0 | | | 10.19 | | -1215000 | 7034 |
| Trunk Lean Mass (g) | | 98.22 | 95.62 | | | -8.583 | 1185000 | 177,699 | -1,711 | -18.5 | 133.2 | 66.8 | | | -9.029 | 742060 | 164848 | -2740 |
| Legs Fat Mass (g) | -7.721 | | 191.1 | | | 3.698 | -1105000 | | -4,432 | -9.0 | -26.4 | 254.0 | | | 4.913 | -1565000 | 489408 | -3014 |
| Legs Lean Mass (g) | -59.64 | 95.58 | 79.11 | | | -8.686 | | 977,288 | -2,272 | -45.7 | 90.0 | 72.2 | | | -5.684 | | 849490 | -3812 |
| Android Fat Mass (g) | 18.13 | -49.07 | 105.8 | | | 3.472 | -209338 | -123,395 | 1,361 | 14.1 | -40.3 | 94.5 | | | 1.992 | | -230232 | 1160 |
| Android Lean Mass (g) | 4.121 | 14.09 | 17.76 | | | -1.014 | 182555 | 40,903 | -684.2 | -1.7 | 20.4 | 12.2 | | | -1.114 | 132212 | 49046 | -962.4 |
| Gynoid Fat Mass (g) | -3.794 | -22.78 | 104.4 | | | 2.871 | -449004 | | -91.79 | | -14.2 | 125.3 | | | 1.665 | -596713 | | -506.6 |
| Gynoid Lean Mass (g) | -21.75 | 23.7 | 32.04 | | | | 409001 | 358,054 | -1,371 | -16.5 | 40.9 | 24.1 | | | -2.138 | 262216 | 186851 | -1513 |
| VAT (g) | 24.74 | -40.6 | 63.86 | | | 2.808 | | -84,386 | 888.4 | 13.6 | -26.2 | 36.2 | | | 1.362 | 116471 | -72967 | 686.8 |
| SCAT (g) | -6.612 | -13.23 | 42.59 | | | 1.379 | -182069 | | 464.2 | | -16.9 | 58.9 | | | 0.916 | -90643 | -137031 | 517.1 |

| | Men: Model C | | | | | | | | | Women: Model C | | | | | | | | |
|---|---|---|---|---|---|---|---|---|---|---|---|---|---|---|---|---|---|---|
| | Age | height | weight | waist | hip | Impedance | Ht2ImLegs | Ht2ImArms | Constant | Age | height | weight | waist | hip | impedance | Ht2ImLegs | Ht2ImArms | Constant |
| Total Fat Mass (g) | 48.98 | -184.9 | 590 | 153.1 | 71.28 | 21.7 | -1881000 | -467,222 | -15,486 | 72.9 | -228.2 | 741.2 | 26.46 | 80.41 | 15.75 | -1556000 | -808571 | -2833 |
| Total Lean Mass (g) | -56.85 | 165.4 | 384.7 | -149.5 | -67.21 | -22.86 | 1751000 | 493,750 | 18759 | -70.2 | 223.4 | 240.6 | -27.49 | -72.34 | -17.08 | 1325000 | 777813 | 3634 |
| Arms Fat Mass (g) | 7.201 | -22.13 | 58.86 | 3.969 | | 2.33 | -157475 | -63,248 | 514.1 | 12.3 | -22.9 | 82.8 | 2.453 | -15.5 | 1.303 | -121058 | -172788 | 2004 |
| Arms Lean Mass (g) | -15.47 | | 62.5 | -18.93 | -31.58 | -3.742 | 929870 | -388,873 | 6,738 | -7.9 | | 34.5 | 2.008 | -19.87 | -1.196 | 724356 | -118528 | 2479 |
| Trunk Fat Mass (g) | 40.69 | -149.3 | 383.3 | 167.6 | -61.27 | 13.65 | -895692 | -394,302 | -3,730 | 54.2 | -167.9 | 373.1 | 162.2 | -38.9 | 11.6 | | -639086 | -2637 |
| Trunk Lean Mass (g) | 11.38 | 78.31 | 190.7 | -75.94 | -45.26 | -9.647 | 907452 | | 8,429 | -16.6 | 138.8 | 107.0 | -11.07 | -39.53 | -10.62 | 508601 | | 1159 |
| Legs Fat Mass (g) | | -14.31 | 142.8 | -18.06 | 131 | 5.879 | -821818 | | -12,645 | 6.4 | -45.5 | 280.2 | -139.2 | 138.2 | 3.893 | -1255000 | | -2910 |
| Legs Lean Mass (g) | -50.28 | 73.39 | 121.1 | -51.15 | 12.93 | -7.665 | | 894,978 | 1,213 | -43.4 | 83.1 | 95.3 | -18.09 | -8.699 | -5.637 | | 787576 | -1716 |
| Android Fat Mass (g) | 9.818 | -39.74 | 72.16 | 44.06 | -17.06 | 4.097 | -113776 | | -1,202 | 9.4 | -31.9 | 64.6 | 39.25 | -8.387 | 2.319 | | -91158 | -1256 |
| Android Lean Mass (g) | 4.559 | 13.44 | 20.14 | -2.501 | | -1.036 | 176399 | 35,143 | -467.8 | -2.0 | 21.0 | 11.6 | 3.346 | -3.271 | -1.108 | 123640 | 58966 | -967.9 |
| Gynoid Fat Mass (g) | | -17.55 | 73.56 | -3.929 | 69.97 | 2.428 | -370996 | -63622 | -5153 | 4.2 | -22.0 | 106.8 | -49.04 | 85.86 | 1.917 | -330774 | -128825 | -3312 |
| Gynoid Lean Mass (g) | -13.52 | 20.66 | 49.85 | -38.19 | 37.53 | -1.2 | 317231 | 225,457 | -881 | -14.5 | 37.6 | 30.5 | -17.43 | 13.01 | -2.21 | 294440 | 129446 | -1173 |
| VAT (g) | 16.66 | -27.53 | 42.74 | 38.98 | -31.76 | 2.463 | | | -96.22 | 10.7 | -20.0 | 26.8 | 26.15 | -19.8 | 1.335 | 58501 | | 177.7 |
| SCAT (g) | -7.054 | -10.09 | 31.14 | 5.799 | 12.07 | 1.281 | -156425 | | -984 | -1.2 | -15.8 | 38.0 | 12.82 | 12.01 | 1.434 | | -71416 | -1463 |
